# One-year outcomes after mass transfers during the first COVID-19 epidemic wave in France. A cohort study

**DOI:** 10.64898/2026.11.25.26368945

**Authors:** Olivier Grimaud, Sylvie Martin, Agnès Ricard-Hibon, Julien Pottecher, Jean-Marc Philippe, Emmanuelle Leray, Sahar Bayat, François Braun, Pierre Carli, Pierre Tattevin, Marc Noizet, Marc-Antoine Sanchez, Manuel Dolz, Albert Vuagnat, Eric Maury, Nicolas Chauliac, Hélène Coignard-Biehler, Nathalie Prieto

**Affiliations:** Univ Rennes, EHESP, CNRS, ARENES – UMR 6051, Rennes, France; Emergency Department SAMU-SMUR 95, CHG Pontoise-Beaumont/Oise, Pontoise, France; Anesthesiology, Critical Care & Perioperative Medicine, Hôpitaux Universitaires de Strasbourg, Strasbourg, France; France Ministry of Health and social Affairs, Directorate General for Health, Paris; CHR Metz-Thionville, Metz, France; Department of Anesthesiology and Intensive Care, SAMU de Paris, Hôpital Necker, Paris, France; Department of Infectious Diseases and Intensive Care Unit, Pontchaillou University Hospital, Rennes, France; Emergency Department and Service d’Aide Médicale Urgente (SAMU) 68, Groupe Hospitalier de la Région Mulhouse Sud Alsace, Mulhouse, France; Information Systems and Digital Department (DSIN), French Army Health Service, Saint-Mandé‑, France; Direction centrale du service de santé des armées, Paris, France; Department for Research, Studies, Evaluation and Statistics (DREES), French Health and Social Affairs Ministry, Paris, France; Université Pierre et Marie Curie, Paris, France; Cellule d’Urgence Médico-Psychologique (CUMP), Centre Régional du Psychotraumatisme (CRP), Hôpital Edouard Herriot, Hospices Civils de Lyon, Lyon, France; RESHAPE (Research on Healthcare Performance), INSERM U1290, Université Claude Bernard - Lyon 1, Lyon, France; Emergency Medical Service, Lyon University Hospital, Lyon, France

**Author notes:** **Corresponding author**, Sylvie Martin, Département METIS (MEthodes quanTItatives en Santé publique), EHESP, 15 Av. du Professeur Léon Bernard, 35043 Rennes.

**Keywords:** ARDS, COVID-19, patient transfer, intensive care units, mortality, psychological distress

## Abstract

**Objectives:** Even taking into account the selection of healthier patients, it has been shown that the mass transfers of patients between intensive care units (ICUs) outside their own region during the first wave of the COVID-19 pandemic in France was associated with lower mortality but a higher rate of psychiatric disorders in the ICU. In other countries, the survival benefit of such transfers had faded after around 60 days. Our aim was to determine the long-term outcomes (up to one year) associated with these mass transfers in France.

**Design:** TRANSCOV retrospective cohort study enriched by data linkage with hospital admissions, outpatient consultations, prescriptions and deaths database.

**Setting:** Multi-centre study involving 39 “origin” ICUs from the East and Paris regions (France) and 57 “destination” ICUs located in regions then unaffected by the pandemic.

**Participants:** Patients with severe COVID-19 (n = 1,113) admitted to ICUs in the Eastern and Paris regions between 4 March 2020 and 10 April 2020.

**Exposure:** Being transferred from versus remaining in overburdened ICUs.

**Outcome measures:** The main outcome was death occurring during the year following admission in ICU. Secondary outcomes related to the mental health of participants during follow-up including hospital admission with a psychiatric disorder as the main or associated diagnosis, outpatient psychiatric consultation and psychotropic drugs prescription.

**Results:** At initial admission, patients in the transfer (n=319) and the control (n=794) groups were similar in age (64 years), sex (74% male), number of comorbidities and severity. However, transferred patients were lighter (-8kg, p<0.001). Respectively 44 and 309 deaths were recorded in the transfer and control groups, the vast majority of which occurring while in ICU. At the end of follow-up, the adjusted incidence rate ratio associated with transfer was 0.18 (95% CI:0.13-0.24) thus suggesting a five-fold lower mortality risk. Transferred patients experienced twice as many psychiatric disorders during initial ICU stay (39.2% versus 18.3%, p<0.001) and these disorders were more often reported as associated (never as main) diagnosis of subsequent hospitalizations (28.8% versus 22%, p=0.04). However, both groups had similar rates of outpatient psychiatric consultations and prescriptions of psychotropic drugs during follow-up.

**Conclusions:** Mass extra-regional inter ICU transfers organised in France in spring 2020 were associated with both short- and long-term survival benefit. Mass-transfer to alleviate overcrowding should be considered an appropriate response to future sudden and localised increase in the demand for intensive care.

**Strengths and limitations of this study:** Reliance on clinical data from ICU medical records allowed adjustment for important initial clinical characteristics in the comparison between groups.

Data-linkage with hospital admissions, outpatient consultations and prescription database allowed long term and unbiased follow-up of both groups.

Our inability to enrol patients transferred abroad questions the generalisability of our results to this population.

Lack of date of occurrence of psychiatric disorders prevented us to explore formally for these outcomes a potential competing risk of death.

## BACKGROUND

During the first wave of the COVID-19 epidemic wave (spring 2020), mass transfers were organized from overcrowded intensive care units (ICUs) in eastern France and Paris to ICUs in regions and neighboring countries that were not yet affected. Previous reports have shown that these transfers of critically ill and heavily medicated patients were safe and were associated with significantly lower short-term mortality in ICU[1–3]. However, studies from other countries have shown less marked contrasts[4,5] and even a disappearance of the survival benefit after about 60 days[6] thus questioning the long-term benefits of transfers. In addition, mass transfers in France have been associated with longer stay, and about twice the rates of delirium and psychological disorders in ICU. In this context, we aimed to study longer-term outcomes (up to one year) following extra-regional mass transfers from France during spring 2020.

## MATERIALS AND METHODS

### Study Design and Setting

We conducted a retrospective, multicenter cohort study (the TRANSCOV cohort) of patients admitted to ICU for COVID-19 between March 4, 2020, and April 10, 2020, with 39 “origin” ICUs from the East of France and Paris regions and 57 “destination” ICUs in others regions of France.The study followed the Strengthening the Reporting of Observational Studies in Epidemiology (STROBE) reporting guidelines for cohort studies

### Data Source

We used individual clinical data extracted from the ICU health records from the TRANSCOV cohort[7]. These data included sociodemographic characteristics, risk factors, comorbidities, severity at admission as well as the dates of the ICU admission, and transfer (for the relevant patients). To extend follow-up, the cohort data were enriched through deterministic data linkage with the French National Health Data System (SNDS). The SNDS database includes records for all hospital admissions, outpatient consultations, prescriptions and, when applicable, the date of death.

### Study Participants

Participants were COVID-19 patients transferred from “origin” ICUs exposed to a sudden overload of patients in March and April 2020 to “destination” ICUs located in areas then unaffected by the pandemic between March 18, 2020, and April 10, 2020. Out of the 502 transferred patients registered in the clinical database, 183 were excluded because of linkage failure or incomplete data (Fig. 1: Study flowchart). The majority of transferred patients with incomplete data had been evacuated abroad (136/144; 94%). Out of a total of 1 280 patients in the clinical database who remained in their “origin” ICU, 1 160 (91%) had complete data. In order to limit indication bias, we aimed to involve only “healthier” control patients who could have been eligible for transfer. Thus, for any transferred patient, one or several controls were selected if they were admitted at the same origin ICU, during the same period (± 2 days), and were alive in ICU at the time of transfer. Based on these criteria, 366 patients were excluded from the control group. The final sample included respectively 319 and 794 patients in the transferred and control groups.

**Fig. 1:**
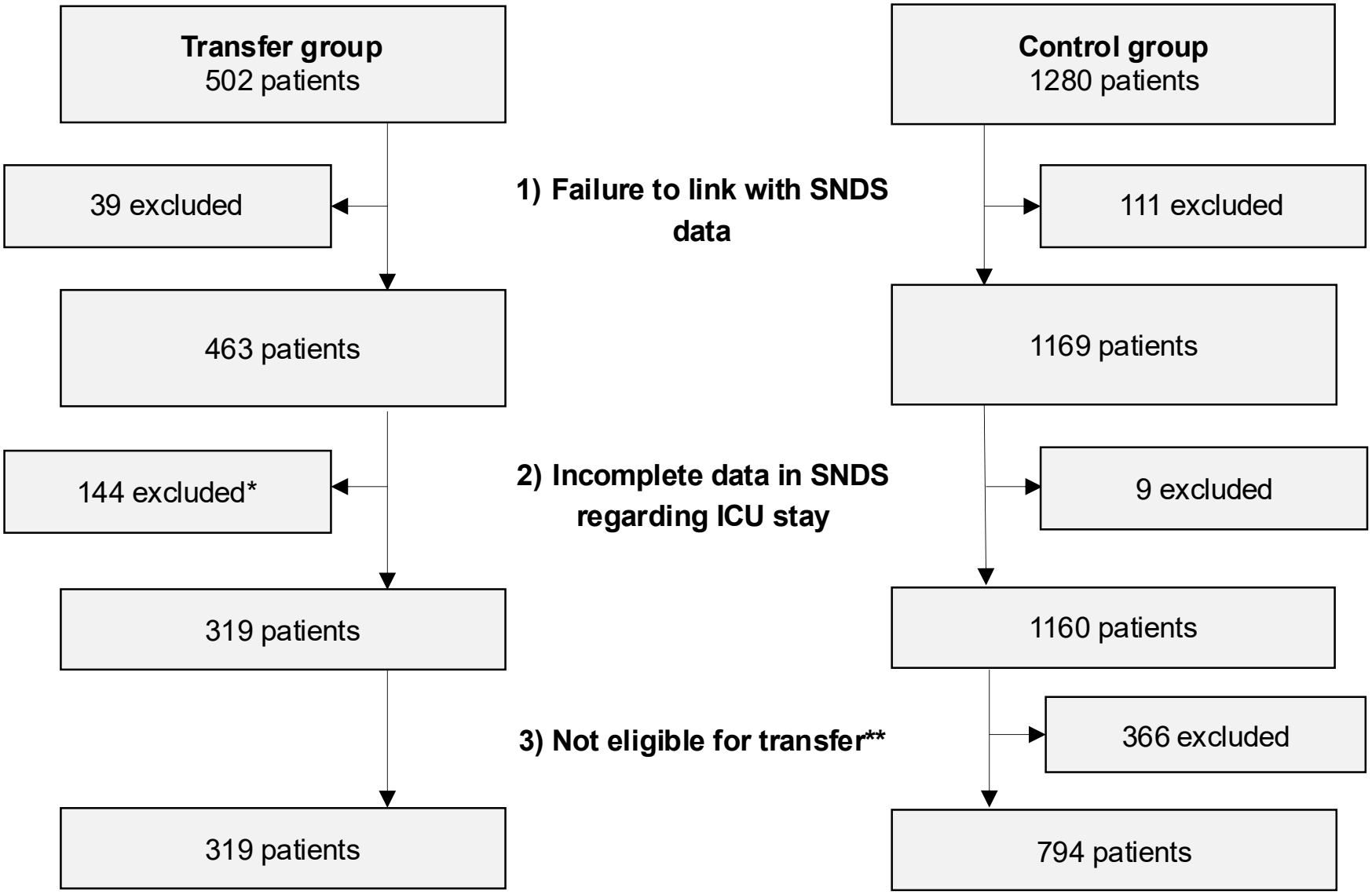
Study flowchart. *of which 136 were transferred abroad **Eligible if control patients matched to index transferred patients using the following criteria: same origin hospital, same ICU admission date (±2 days), present and alive in ICU on the transfer date

### Patient and public involvement

None.

### Outcomes

The main outcome was survival up to one year after ICU admission. The secondary outcome is the identification of psychiatric disorders as primary or secondary diagnoses during new hospital admissions (acute and non-acute care) during the year following hospital discharge. We also identified outpatient visits to psychiatrists and psychotropic medication. The latter were defined as positive if at least three prescriptions were dispensed within one year after hospital discharge (codes in eAppendix 1 in the supplement).

### Statistical Analysis

We used Wilcoxon-Mann-Whitney tests for quantitative variables and Chi-square or Fisher’s exact tests for qualitative variables. We plotted Kaplan-Meïer survival curve and compared incidence of deaths and of other clinical events between the groups using mixed Poisson regression models. The models included a random effect at the level of origin ICU and the log of follow-up time as an offset. Co-variables for calculating adjusted associations included: age, number of comorbidities (0,1 and >=2) and Simplified Acute Physiology Score (SAPS II) (in tertiles: <37, 37-49 and >49) at initial admission. R Studio version 4.1.0 was used for analyses. Statistical significance was defined as a two-sided P-value < 0.05.

## RESULTS

At initial admission, the 319 transferred and 794 control patients were similar in age, sex, comorbidity and physiological score, but the former had significantly lower body weight (median: - 8 kg) (table 1). Most deaths in acute care occurred while in ICU. At the end of acute care, the risk in the transferred group was significantly lower: 12.9% versus 38.2%, with an adjusted incidence rate ratio (aIRR) of 0.23 (95%CI:0.17-0.32). Transferred patients remained longer in acute care (+ 8 days), mostly because of longer stay in ICU (+ 6 days). During the initial hospital stay, both groups experienced comparable rates of acute renal failure and of nosocomial infections. However, psychiatric disorders were overall twice as common among transferred patients (39.2% versus 18.3%, p<0.001). Given their much higher risk of ICU mortality, patients in the control group who were likely to have psychiatric disorders may have been excluded from the analysis due to early mortality, thereby reducing the frequency of psychiatric disorders. As our data does not specify the date on which the clinical events occurred, we were unable to formally investigate this hypothesis of competing risks. However, the association remained positive and significant once taking account of length of stay in adjusted mixed Poisson model (aIRR of psychiatric disorders transfer/control =1.66, 95%CI:1.30-2.12). Similar results applied in analysis including only patients who survived ICU (aIRR=1.68, 95%CI:1.30-2.18). Neurotic, stress and somatoform disorders, followed by mood [affective] disorders were the two most frequent diagnostic categories, and about twice as common in the transferred group.

**Table 1:**
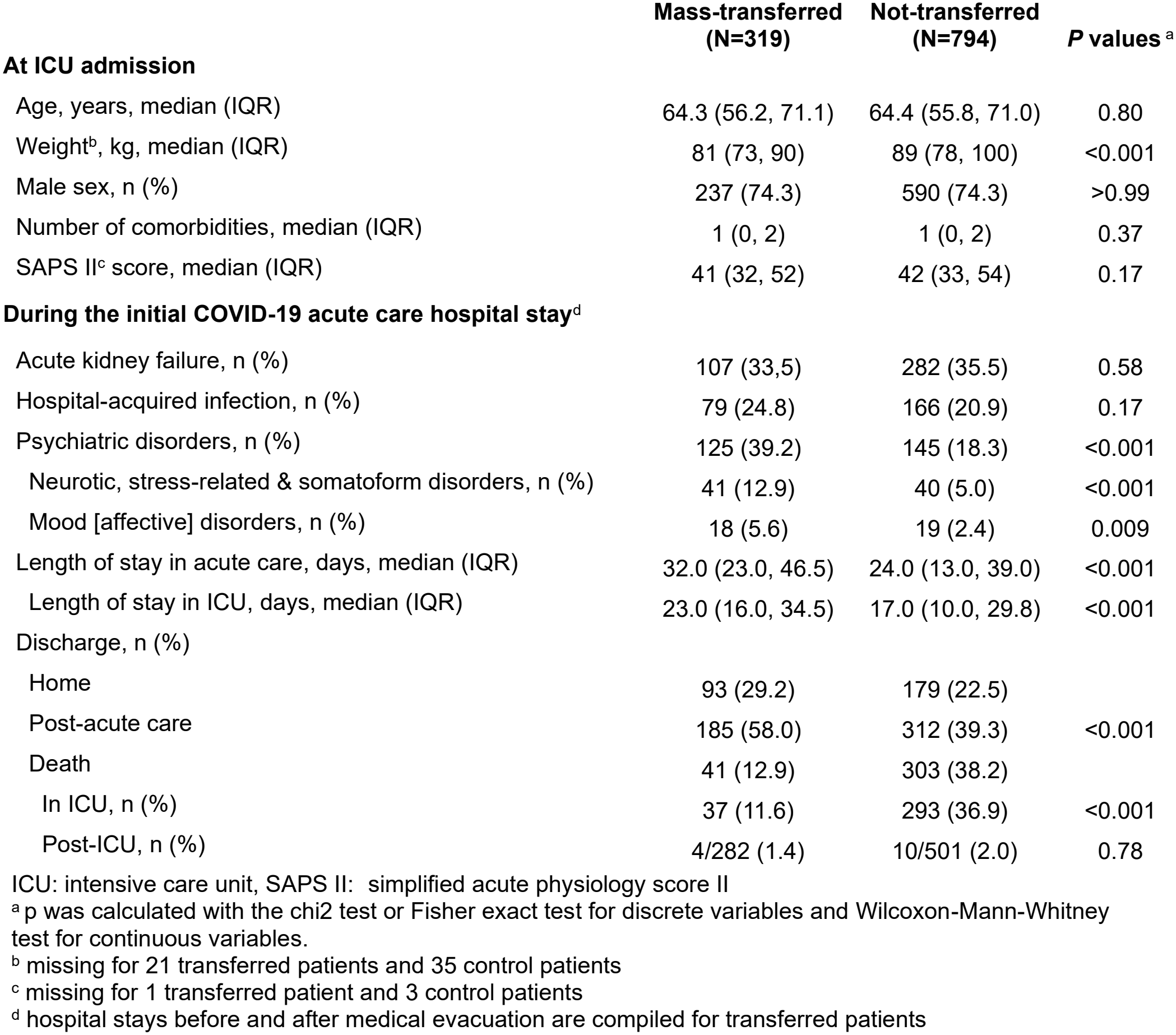
Clinical characteristics and outcomes during initial hospital stay.

|  | **Mass-transferred (N=319)** | **Not-transferred (N=794)** | ***P* values** <sup>a</sup> |
|---|---|---|---|
| **At ICU admission** |  |  |  |
| Age, years, median (IQR) | 64.3 (56.2, 71.1) | 64.4 (55.8, 71.0) | 0.80 |
| Weight<sup>b</sup>, kg, median (IQR) | 81 (73, 90) | 89 (78, 100) | <0.001 |
| Male sex, n (%) | 237 (74.3) | 590 (74.3) | >0.99 |
| Number of comorbidities, median (IQR) | 1 (0, 2) | 1 (0, 2) | 0.37 |
| SAPS II<sup>c</sup> score, median (IQR) | 41 (32, 52) | 42 (33, 54) | 0.17 |
| **During the initial COVID-19 acute care hospital stay**<sup>d</sup> |  |  |  |
| Acute kidney failure, n (%) | 107 (33,5) | 282 (35.5) | 0.58 |
| Hospital-acquired infection, n (%) | 79 (24.8) | 166 (20.9) | 0.17 |
| Psychiatric disorders, n (%) | 125 (39.2) | 145 (18.3) | <0.001 |
| Neurotic, stress-related & somatoform disorders, n (%) | 41 (12.9) | 40 (5.0) | <0.001 |
| Mood [affective] disorders, n (%) | 18 (5.6) | 19 (2.4) | 0.009 |
| Length of stay in acute care, days, median (IQR) | 32.0 (23.0, 46.5) | 24.0 (13.0, 39.0) | <0.001 |
| Length of stay in ICU, days, median (IQR) | 23.0 (16.0, 34.5) | 17.0 (10.0, 29.8) | <0.001 |
| Discharge, n (%) |  |  |  |
| Home | 93 (29.2) | 179 (22.5) |  |
| Post-acute care | 185 (58.0) | 312 (39.3) | <0.001 |
| Death | 41 (12.9) | 303 (38.2) |  |
| In ICU, n (%) | 37 (11.6) | 293 (36.9) | <0.001 |
| Post-ICU, n (%) | 4/282 (1.4) | 10/501 (2.0) | 0.78 |

ICU: intensive care unit, SAPS II:  simplified acute physiology score II
<sup>a</sup> p was calculated with the chi2 test or Fisher exact test for discrete variables and Wilcoxon-Mann-Whitney test for continuous variables.
<sup>b</sup> missing for 21 transferred patients and 35 control patients
<sup>c</sup> missing for 1 transferred patient and 3 control patients
<sup>d</sup> hospital stays before and after medical evacuation are compiled for transferred patients

Outcomes during the 12-month follow-up are shown in table 2. Mortality was then low and similar in both groups (1.4%), so that the survival benefit of transfer at the end of the initial hospital admission was sustained throughout the follow-up period (see Fig. 2). Regarding one-year survival following admission to the ICU, the aIRR of 0.18 (95%CI:0.13-0.24) translates into a five times lower mortality among the transferred compared to the control group (crude IRR= 0.19 (95%CI:0.14-0.27)). Fewer than 1% of patients were admitted to a psychiatric hospital during the follow-up period. However, psychiatric disorders were more frequently listed among the associated diagnoses in transferred patients during other hospital admissions (28.8% versus 22.0%, p = 0.04). Although neurotic, stress and somatoform disorders remained the most frequent diagnostic category, mood disorders frequency were higher during follow-up and again twice as frequent in the transferred group. In both groups, less than one in ten patients benefited from psychiatric consultations, and one in four received at least three dispensing of psychotropic drugs during follow-up. The overall pattern of psychotropic drugs prescriptions did not differ significantly between groups. Patients who experienced psychological distress during their ICU stay were more likely to seek psychiatric consultations (12.9% versus 5.9%, p=0.002) or to use psychotropic drugs (35.9% versus 19.5%, p<0.001). This pattern applied similarly in the transferred and control groups.

**Table 2:** Mortality and psychiatric outcomes during the 12 months after discharge from initial COVID-19 acute hospital care.

|  | **Mass-transferred (N=278)** | **Not-transferred (N=491)** | ***P* values** <sup>a</sup> |
|---|---|---|---|
| **Mortality** |  |  |  |
| Death during the 12 months follow-up, n (%) | 4 (1.4) | 7 (1.4) | >0.99 |
| **Psychiatric outcomes** |  |  |  |
| Admission to psychiatric hospital, n (%) | 1 (0.4) | 4 (0.8) | 0.66 |
| Admission in other hospitals with psychiatric code<br>as associated diagnosis<sup>b</sup>, n (%) | 80 (28.8) | 108 (22.0) | 0.04 |
| Neurotic, stress-related & somatoform disorders, n (%) | 34 (12.2) | 41 (8.4) | 0.01 |
| Mood [affective] disorders, n (%) | 29 (10.4) | 24 (4.9) | 0.005 |
| Psychiatrist outpatient (with one or more), n (%) | 18 (6.5) | 42 (8.6) | 0.33 |
| Number per patient, median (IQR) | 3.0 (1.3, 9.5) | 2.0 (1.0, 6.0) | 0.45 |
| Psychotropic drugs use<sup>c</sup>, n (%) | 68 (24.5) | 116 (23.6) | 0.79 |
| Anxiolytic and hypnotic<sup>c</sup>, n (%) | 51/68 (75.0) | 80/116 (69.0) | 0.40 |
| Antidepressant<sup>c</sup>, n (%) | 35/68 (51.5) | 60/116 (51.7) | >0.99 |
| Others<sup>c</sup>, n (%) | 4/68 (5.9) | 14/116 (12.1) | 0.21 |

<sup>a</sup> p was calculated with the chi2 test or Fisher exact test for discrete variables and Wilcoxon-Mann-Whitney test for continuous variables.
<sup>b</sup> there were no psychiatric code recorded as main diagnosis.
<sup>c</sup> with three pharmaceutical dispensing or more during the year following hospital discharge.

**Fig. 2:**
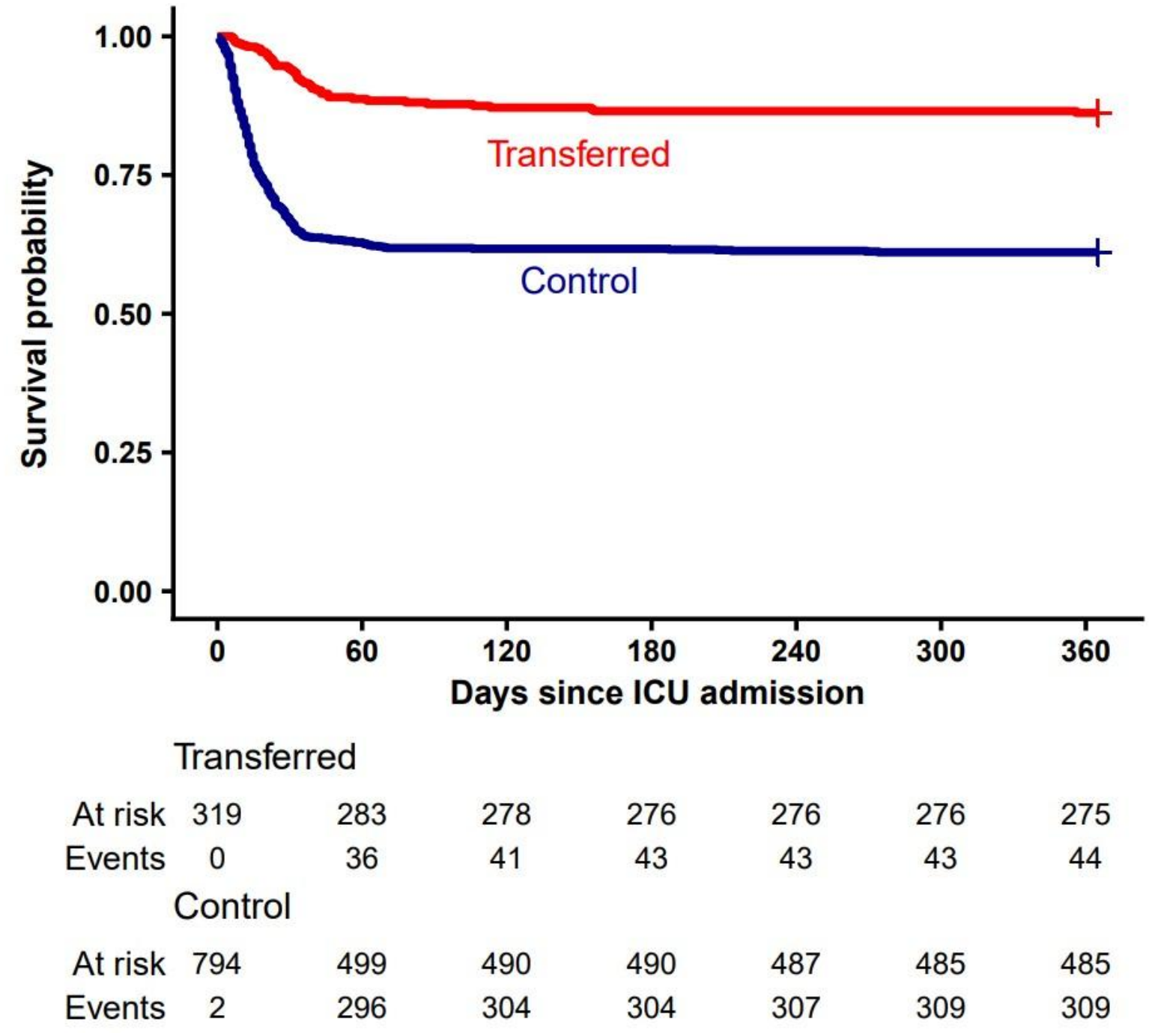
Kaplan-Meïer one-year survival curve.

## DISCUSSION

Our main and important finding is that the considerably lower risk of short-term mortality associated with mass transfers of spring 2020 in France is sustained up to one year. The approximately fourfold reduction in mortality risk (1/0.23) observed at the end of the initial acute care stay in the transfer group is consistent with previous findings[8]. Over the following year, both groups showed low and comparable levels of risk, which resulted in a comparable benefit in terms of one-year survival (five times lower mortality rate) associated with the transfer. This contrasts with results obtained elsewhere, notably those from a national study in the Netherlands which identified a marginal and transient survival benefit (up to 60 days) associated with transfer[6]. In this study transferred patients were compared to those from all other ICUs in the national territory, thus averaging the burden of care to which transferred and control patients were exposed. Our decision to select patients for the control group from those who had remained in their original intensive care unit — which was under considerable strain at the time — likely helped to highlight the benefits of being transferred out of an overburdened care setting[8]. Apart from differences in design, comparison with other studies may also be hampered by varying study periods (sometimes covering several epidemic waves), epidemic dynamic, geographical and health care contexts. A constant result though is the longer length of stay in ICU associated with transfer (plus 8 days[5], plus 15 days[4]).

To our knowledge, the TRANSCOV study is the only one reporting higher (∼twice) frequency of psychiatric disorders in the transfer group during initial hospitalization. This may be because patients received more attention and care in less crowded ICUs following their transfer. It could also be the consequence of a competing risk exerted by early death among susceptible patients in the control group. However, our analysis does not appear to support this hypothesis as the sole explanation for our findings. An alternative explanation is that the transfer process itself and, perhaps, remoteness from relatives after medical evacuation exacerbated stress. Based on psychiatric outpatients and psychotropic drugs prescriptions, we found however no evidence of clinically important medium-term mental health consequences associated with transfer. This finding adds and agrees with those of two small-scale qualitative studies which found no long-term (>12 months) impact of transfers[9,10].

A limit of our analysis is that it involved about half of all patients transferred extra-regionally in spring 2020 in France (319 over an estimated 660). This is mainly due to inability to enrol patients transferred abroad and, to a lesser extent, to failure of linkage between clinical and SNDS data. Lack of date of event in the clinical database prevented to explore more thoroughly the competing risk hypothesis. Furthermore, the SNDS does not provide any information on quality-of-life outcomes, which may have varied between groups. Finally, despite our efforts to limit indication bias, part of the survival benefit associated with transfer may still stem from the fact that clinical team evacuated patients with better prognosis.

## CONCLUSION

Overall, our study adds to the body of evidence suggesting that organizing mass transfers between ICU is a safe and viable option for managing a sudden, acute and localized influx of critically ill patients.

## Supporting information

STROBE

Supplemental eAppendix1

## DECLARATIONS

## Acknowledgements

The authors would like to thank the TRANSCOV collaborators who helped collect the clinical data. The Centre of Clinical Investigation of the Rennes teaching hospital was involved in setting up and managing the clinical database. The Health Data Hub provided technical assistance for linking clinical data to the SNDS database.

## Contributors

OG had full access to all of the data in the study and takes responsibility for the integrity of the data and the accuracy of the data analysis.

Concept and design: OG, EL, SB.

Acquisition, analysis, or interpretation of data: all authors. Drafting of the manuscript: OG, SM.

Critical review of the manuscript for important intellectual content: all authors. Statistical analysis: SM.

Obtained funding: OG.

Administrative, technical, or material support: OG. Supervision: OG.

### Funding

The study was funded by the Directorate General of Health (DGS) of the French Ministry of Health and received supplementary funding from the French agency for AIDS and viral hepatitis research, emerging infectious diseases (ANRS MIE). The TRANSCOV cohort has been labeled as a National Research Priority by the National Orientation Committee for Therapeutic Trials and other research on COVID-19 (CAPNET).

### Disclaimer

The funders had no role in the design and conduct of the study; collection, management, analysis, and interpretation of the data; preparation, review, or approval of the manuscript; and decision to submit the manuscript for publication.

### Patient and public involvement

Patients and/or the public were not involved in the design, or conduct, or reporting, or dissemination plans of this research.

### Patient consent for publication

Not applicable.

### Ethics approval

The study protocol was approved by the French national ethics and scientific committee on health research (CESREES, decision n° 2046524, 16/07/2020) and by the French data protection authority (CNIL, decision n° DR-2022-234, 8/11/2022). All included patients received a document with information on the study objectives and methods as well as explanations on the right and procedures to decline participation.

### Provenance and peer review

Not commissioned; externally peer reviewed

### Data availability statement

According to data protection and the French regulation, the authors cannot publicly release the data from the French national health data system (SNDS). However, a request for data reuse may be made and would require prior approval from the French regulatory authorities.

