## Supplementary material for "One-year outcomes after mass transfers during the first COVID-19 epidemic wave in France. A cohort study": STROBE

**STROBE-checklist-TRANSCOV**

STROBE Statement—Checklist of items that should be included in reports of ***cohort studies***

|  | Item No | Recommendation | Check |
| --- | --- | --- | --- |
| **Title and abstract** | 1 | (*a*) Indicate the study’s design with a commonly used term in the title or the abstract | The title includes the word “Cohort” |
|  |  | (*b*) Provide in the abstract an informative and balanced summary of what was done and what was found | We focussed on the specific design of our study (most importantly the data-linkage process which enabled follow-up). We also reported results regarding the main (survival) and secondary outcomes. |
| Introduction | | |  |
| Background/rationale | 2 | Explain the scientific background and rationale for the investigation being reported | Preceding reports investigating the same event are quoted and reviewed and the rationale for conducted the specific analysis addressed. |
| Objectives | 3 | State specific objectives, including any prespecified hypotheses | The introduction ends with the statement of the aim of the study. |
| Methods | | |  |
| Study design | 4 | Present key elements of study design early in the paper | The materials and methods’ section includes details regarding the design: multicentre cohort enriched via data-linkage with medico-administrative database enabling follow-up. The participants include COVID-19 patients transferred in spring 2020 from the East and Paris regions of France towards other regions. Those are compared to a subset of “non-transferred” patients included in the cohort and who could have been, according to available information, eligible for medical evacuation. |
| Setting | 5 | Describe the setting, locations, and relevant dates, including periods of recruitment, exposure, follow-up, and data collection |  |
| Participants | 6 | (*a*) Give the eligibility criteria, and the sources and methods of selection of participants. Describe methods of follow-up |  |
|  |  | (*b*) For matched studies, give matching criteria and number of exposed and unexposed |  |
| Variables | 7 | Clearly define all outcomes, exposures, predictors, potential confounders, and effect modifiers. Give diagnostic criteria, if applicable | The “data source” paragraph describes date, sources and the variables collected. Exposed (transferred patients) and non-exposed (control patients) were assessed in a similar manner by extracting information from ICU medical records and French National Health Data System database for a one-year follow-up period. |
| Data sources/ measurement | 8* | For each variable of interest, give sources of data and details of methods of assessment (measurement). Describe comparability of assessment methods if there is more than one group |  |
| Bias | 9 | Describe any efforts to address potential sources of bias | By selecting for each transferred patient, control(s) who were admitted in the same ICU at the same period and were still alive at the date of transfer allowed to avoid immortal time bias and to control for contextual factors. |
| Study size | 10 | Explain how the study size was arrived at | As explained in the companion protocol paper the Transcov cohort is well powered (99%) for detecting a 10% risk difference in mortality outcome. In this specific analysis of this paper where 319 transferred patients are matched to 794 controls, the statistical power to detect a 10% difference in ICU mortality is 93%. |
| Quantitative variables | 11 | Explain how quantitative variables were handled in the analyses. If applicable, describe which groupings were chosen and why | The number of comorbidities and the SAPS II score were categorized to address the lack of log-linearity in the models (Statistical analysis paragraph). |
| Statistical methods | 12 | (*a*) Describe all statistical methods, including those used to control for confounding | See paragraphs “Study Participants” and “statistical analysis”. |
|  |  | (*b*) Describe any methods used to examine subgroups and interactions | Given the small number of deaths occurring after discharge from the initial COVID-19 admission (which is the specific study period being focussed on in this paper), we did not perform subgroup analysis for this outcome. Regarding psychiatric disorders, we examined whether occurrence of such troubles during initial ICU stay was associated with therapeutical events during follow-up. See also 1rst parag of the results section where modelling psychiatric disorders occurrence excluding patients who died in ICU is presented. |
|  |  | (*c*) Explain how missing data were addressed | Not applicable: only 4 patients had missing data among the variables used in the models. |
|  |  | (*d*) If applicable, explain how loss to follow-up was addressed | The analysis study includes only patients whose clinical data from the TRANSCOV cohort were linked to the French National Health Data System database. This database is virtually exhaustive regarding vital status. |
|  |  | (*e*) Describe any sensitivity analyses | Not applicable for the main outcome (survival) given the few numbers of events after discharge from initial COVID-19 admission (main focus of this paper). |
| Results | | |  |
| Participants | 13* | (a) Report numbers of individuals at each stage of study—eg numbers potentially eligible, examined for eligibility, confirmed eligible, included in the study, completing follow-up, and analysed | See the flow diagram (fig 1) and corresponding text at the beginning of the results section |
|  |  | (b) Give reasons for non-participation at each stage |  |
|  |  | (c) Consider use of a flow diagram |  |
| Descriptive data | 14* | (a) Give characteristics of study participants (eg demographic, clinical, social) and information on exposures and potential confounders | Table 1 summarises demographic and clinical characteristics of the transferred and control patients. |
|  |  | (b) Indicate number of participants with missing data for each variable of interest | Yes, this is explained sequentially in the paper. Missing values did not exceed 5%. |
|  |  | (c) Summarise follow-up time (eg, average and total amount) | We present Kaplan Meier graphs with numbers of patients still at risk at events every 2 months. |
| Outcome data | 15* | Report numbers of outcome events or summary measures over time | See Kaplan Meier graphs for the main outcomes. |
| Main results | 16 | (*a*) Give unadjusted estimates and, if applicable, confounder-adjusted estimates and their precision (eg, 95% confidence interval). Make clear which confounders were adjusted for and why they were included | Regarding survival, crude and adjusted incidence rate ratio are presented with 95% confidence intervals in the text of the results section. The co-variables used in adjusted models are presented in the method section. |
|  |  | (*b*) Report category boundaries when continuous variables were categorized | Yes, see paragraph “statistical analysis” |
|  |  | (*c*) If relevant, consider translating estimates of relative risk into absolute risk for a meaningful time period | N/A |
| Other analyses | 17 | Report other analyses done—eg analyses of subgroups and interactions, and sensitivity analyses | Yes, See also answer above in Statistical methods |
| Discussion | | |  |
| Key results | 18 | Summarise key results with reference to study objectives | See first paragraph of the discussion |
| Limitations | 19 | Discuss limitations of the study, taking into account sources of potential bias or imprecision. Discuss both direction and magnitude of any potential bias | Limitations are addressed in the last paragraph of the discussion. The inability to involve patients transferred abroad is specifically discussed. We endeavoured to formulate interpretation building on previous reports and of the relevant scientific literature. |
| Interpretation | 20 | Give a cautious overall interpretation of results considering objectives, limitations, multiplicity of analyses, results from similar studies, and other relevant evidence |  |
| Generalisability | 21 | Discuss the generalisability (external validity) of the study results | The paper describes an exceptional situation, however lessons can be drawn and may prove useful for circumstances where ICU are locally overwhelmed, as suggested in the conclusion. |
| Other information | | |  |
| Funding | 22 | Give the source of funding and the role of the funders for the present study and, if applicable, for the original study on which the present article is based | As requested by the journal |

*Give information separately for exposed and unexposed groups.

**Note:** An Explanation and Elaboration article discusses each checklist item and gives methodological background and published examples of transparent reporting. The STROBE checklist is best used in conjunction with this article (freely available on the Web sites of PLoS Medicine at http://www.plosmedicine.org/, Annals of Internal Medicine at http://www.annals.org/, and Epidemiology at http://www.epidem.com/). Information on the STROBE Initiative is available at http://www.strobe-statement.org.
