## Supplemental eAppendix1 for "One-year outcomes after mass transfers during the first COVID-19 epidemic wave in France. A cohort study"

**Supplemental Content**

**eAppendix 1: Variables extracted from the SNDS tables with their codes used to construct the indicators**

**At admission**

- **Number of comorbidities:** Number of comorbidities identified in the medical records from the following list: treated diabetes, treated arterial hypertension, treated ischemic heart disease, stroke within the last 12 months, treated heart failure, renal failure requiring dialysis, COPD, treated asthma, cancer under treatment or active within the last 12 months, immunosuppression.
- **SAPS II score:** the highest SAPS II score between the values reported in the medical records and the score in the variable *IGS2_COD* from the *T_MCOaaUM* table (in the SNDS) for the ICU stay.

**During ICU or acute care stays**

- **Acute kidney failure:** ICD‑10 = *N17* in variables *DGN_PAL* or *DGN_REL* of tables *T_MCOaaB* and *MCOaaUM*, or *ASS_DIAG* of table *T_MCOaaD*.
- **Hospital‑acquired infection:** ICD‑10 = *Y95* and ICD‑10 codes starting with *A*, *B*, *J0*, *J1* or *J2* in variables *DGN_PAL* or *DGN_REL* of tables *T_MCOaaB* and *MCOaaUM*, or *ASS_DIAG* of table *T_MCOaaD*.
- **Psychiatric disorders:** ICD‑10 codes starting with *F* in variables *DGN_PAL* or *DGN_REL* of tables *T_MCOaaB* and *MCOaaUM*, or *ASS_DIAG* of table *T_MCOaaD*.

**After the acute care stay**

- **Psychiatric disorders in acute and post-acute care:**

ICD‑10 codes starting with *F*

- in variables *DGN_PAL* or *DGN_REL* of tables *T_MCOaaB* and *MCOaaUM*, or *ASS_DIAG* of table *T_MCOaaD*, or
- in variables *MOR_PRP* or *ETL_AFF* of table *T_SSRaaB*, or *DGN_COD* of table *T_SSRaaD*, or
- **Admission to psychiatric hospital:**

ICD‑10 codes starting with *F* in variable *DGN_PAL* of table *T_RIPaaRSA*

- **Psychiatric consultation:**

1. in table *ER_PRS_F*,
   - variable *PSE_SPE_COD* = 17 or 33 or 75, and
   - at least one of the variables *PRS_NAT_REF*, *BSE_PRS_NAT*, *CPL_PRS_NAT* contains one of the following procedure codes:
     1102, 1113, 1118, 1216, 2247, 2380–2389,
     **or**
2. in table MCO (*T_MCOaaFCSTC*) or table SSR (*T_SSRaaFCSTC*),
   - variable *EXE_SPE* = 17 or 33 or 75, and
   - at least one of the variables *PRS_NAT_REF*, *BSE_PRS_NAT*, *CPL_PRS_NAT* contains one of the following procedure codes:
     APY, AVY, CNP, CNPSY, VNPSY, MCY, 2TC, TCS 2, MCPCNPSY, FSY, PY0–PY9,
     **or**
3. in the psychiatric consultation table *T_RIPaaR3A*,
   - all consultations.

- **Psychotropic drugs dispensing:**

ATC codes = *N05* or *N06* in variable *PHA_ATC_CLA* of table *IR_PHA_R*, dispensed at least 3 times in the year following discharge from acute care.

- **Anxiolytic and hypnotic drugs dispensing:**

ATC codes = *N05B* or *N05C* in variable *PHA_ATC_CLA* of table *IR_PHA_R*, dispensed at least 3 times in the year following discharge from acute care.

- **Antidepressant drugs dispensing:**

ATC code = *N06A* in variable *PHA_ATC_CLA* of table *IR_PHA_R*, dispensed at least 3 times in the year following discharge from acute care.

- **Other psychotropic drugs dispensing:**

ATC codes = *N05A*, *N06B*, or *N06D* in variable *PHA_ATC_CLA* of table *IR_PHA_R*, dispensed at least 3 times in the year following discharge from acute care.
